# Stability of HIV RNA in plasma stored at different temperatures

**DOI:** 10.64898/2026.09.22.26363748

**Authors:** Molly Levine, Ingrid A. Beck, Rachel A. Silverman, Greg Pepper, Lisa M. Frenkel

**Affiliations:** Center for Global Infectious Disease Research, Seattle Children’s Research Institute, Seattle, Washington, USA; Center for Biostatistics and Health Data Science, Department of Statistics, Virginia Tech, Roanoke, Virginia, USA; Department of Laboratory Medicine and Pathology, University of Washington, Seattle, Washington, USA; Department of Pediatrics, University of Washington, Seattle, Washington, USA

**Author notes:** Corresponding author (LF). co-first authors (Molly Levine, Ingrid A. Beck).

## Abstract

In resource-limited settings plasma is usually transported to centralized laboratories for HIV RNA quantification; often by motor vehicles using dry ice. Any decay in plasma HIV RNA when near the threshold used to define virologic failure could impact clinical management. However, maintaining a cold chain during transport can be costly and limit the frequency of shipments, which can delay clinical assessments and also negatively impact patient care. Transporting plasma at ambient temperatures could facilitate timely measurements of plasma HIV RNA viral loads (**VL**). We conducted a stability study of HIV RNA in plasma held for 1, 3, 5, and 7 days at a range of temperatures (4°C, 30°C, and 37°C) to simulate the durations and temperatures specimens could experience during transport. Thirteen large volume plasma specimens with VL ranging from 2.7-5.53 log_10_ copies/mL were evaluated. Mean changes in VL between Day-0 to Day-1, Day-3, Day-5 or Day-7 were calculated, with decays >0.5 log_10_ c/mL considered clinically relevant. Compared to Day 0, the VL of plasma stored at 4°C and 30°C remained stable across the Days tested. The VL in plasma stored at 37°C had a progressive decline at Day-1, Day-3, Day-5 and Day-7 (means of - 0.04, -0.19, -0.33. -0.34 log_10_ c/mL, respectively), with a decay of >0.5 log_10_ observed in 3/13 (23%) specimens at Day-7. A linear regression analysis showed that plasma specimens stored at 37°C significantly decayed over time (slope - 0.054, 95% CI -0.079, -0.029; p=0.01). Any decay observed among specimens stored at 4°C and 30°C was not clinically relevant nor statistically significant. Our observations suggest that HIV RNA levels in plasma should remain stable if transported to centralized laboratories at ambient temperatures of ≤30°C or chilled by cold packs over a period ≤7 days. Transporting specimens with cold packs instead of dry ice could reduce costs and facilitate the timely monitoring of VL used to assess and guide antiretroviral therapy.

## Introduction

The World Health Organization (**WHO**) recommends that individuals living with human immunodeficiency virus type-1 (**HIV**) who are taking antiretroviral therapy (**ART**) have treatment efficacy monitored by quantification of the plasma HIV RNA viral load (**VL**), and genotypic testing for drug resistance if viral replication is not suppressed by ART after adherence counseling [1]. In resource limited settings, these assays are generally available only in centralized laboratories, requiring the transport of specimens from smaller and sometimes remote clinics to these laboratories.

WHO guidelines recommend blood processing and storage on several timelines and temperatures prior to VL testing [2]. First, whole blood maintained at room temperature should be processed within 6 hours, or if refrigerated between 2°C and 8°C it should be processed within 24 hours. Once plasma is separated, refrigeration between 2°C and 8°C is recommended for a maximum of 5 days. If longer storage periods are required, freezing at -20°C or lower temperatures is recommended.

Alternative specimen types can be transported at ambient temperatures to monitor VL or genotype HIV, including whole blood or plasma dried on filter paper. VL determined from dried blood spots (**DBS**) on Whatman 903 paper provide values comparable to frozen plasma when an individual’s VL is >1000 copies/mL (**c/mL)** [3–7], but results are variable at lower VL [4, 6, 7]. Due to the relatively small sample volume in DBS (50 μL/spot), the efficiency of genotyping is significantly reduced when HIV RNA is <2000 c/mL (e.g., 12/22 successfully genotyped [8]), and the detection of minor frequency populations in an individual’s HIV quasispecies can be limited [3]. Utilization of dried plasma spots and larger capacity systems like ViveST tubes for plasma storage or transport at ambient temperatures lose varying amounts of HIV RNA [9–11]. Recently, the Cobas plasma separation card has shown accurate HIV VL measurement in primary health care settings and stability [12] when sample cards were stored between 25°C and 42°C for up to 28 days [13]; with a limit of detection of ∼800 c/mL in plasma collected on these cards [14]. Thus, frozen storage of plasma is preferred over DBS and other dried systems for both quantification of HIV RNA [15] and drug resistance testing [16].

Dry ice is often used to keep specimens frozen during shipping from distant clinics to central laboratories. In resource limited settings, dry ice may be costly or irregularly available, leading to batched and infrequent shipment of specimens, which can delay testing, including timely detection of virologic failure or selection of drug resistance mutations. Unidentified ART failure can allow drug resistance mutations to accumulate, and increase transmission of HIV due to unsuppressed virus replication [17].

Several studies suggest that HIV RNA is stable in plasma beyond current recommendations for VL testing at 4°C and 25°C [18–22]. However, the stability at higher ambient temperatures is less certain [20, 21] as most of these studies had small sample sizes and did not include samples with low-level viremias (<3,000 c/mL) [22].

Given the relative ease of transporting specimens at ambient temperatures, the preliminary data showing HIV RNA may be stable in plasma at ambient temperatures for ≤7 days, combined with the improved reproducibility of quantifying low-level VL in plasma compared to DBS [6, 15, 16], we aimed to define the stability of HIV RNA across a range of VL, temperatures and time intervals. We reasoned that if HIV RNA in unfrozen plasma remains stable for one week, this would provide data in support of plasma transport to centralized laboratories for VL testing using Styrofoam boxes with cold packs or in mild climates at ambient temperatures; bypassing the need for dry ice or couriers with transport freezers.

## Materials and Methods

### Study design

The stability of HIV RNA held for a week at various temperatures was assessed using de-identified remnant plasma specimens from HIV-infected individuals. The remnant specimens were fully anonymized prior to our use. We used a laboratory developed HIV RNA quantitative real-time PCR (qPCR), as described below, to determine the plasma HIV RNA levels in these anonymized remnant specimens. For this study no clinical data associated with the anonymized remnant plasma specimens were obtained. Use of these plasmas was determined to not involve human subjects by the University of Washington’s Human Subjects Institution Review Committee therefore participant consent was not performed.

The intra- and inter-assay variability of the quantitative HIV RNA qPCR was determined using aliquots of our laboratory’s stock of “control plasma” made from a pool of 20 remnant plasmas. The VL of the control stock was 5.78 log_10_ c/mL (Abbott RealTime HIV-1 VL assay, Abbott Molecular Inc, Des Plains, IL). The control stock was diluted with uninfected plasma to create multiple single-use plasma aliquots with VL of 4.64 (C-h for “high”) and 3.64 (C-l, for “low”) log_10_ c/mL, which were stored frozen for later use. To estimate the intra-assay variation of our HIV qPCR four aliquots of C-h and four of C-l were extracted and tested in a single assay. To determine the inter-assay variation of our qPCR, two aliquots each of C-h and C-l were extracted and tested on two additional dates.

Assay variability was assessed by calculating the mean VL and standard deviation (**SD**) for the C-h and C-l controls.

Plasma HIV RNA stability at various temperatures were evaluated using a set of 13 unique “experimental plasma” specimens (S1-S13, VL range 2.70-5.53 log_10_ c/mL) that were held at 4°C, 30°C, or 37°C for 0 to 7 days. These included three neat plasmas (S3-S5), seven mixtures of two plasmas with similar VL (S1, S2, S6-S10) and three plasmas diluted with uninfected plasma (S11-S13) to create samples with VL near 3 log_10_ (1000) c/mL. To limit the variables tested to plasma storage temperature over time, each experimental specimen was divided into 15 single-use 140 µL aliquots. Two aliquots of each plasma had RNA extracted and reverse transcribed into cDNA immediately (Day 0). This cDNA (which is relatively stable compared to RNA) was then frozen at -20°C for later HIV qPCR VL testing. Four aliquots of the remaining 13 aliquots from each experimental plasma were stored at 4°C, 30°C, or 37°C (four aliquots at each temperature) to evaluate stability at these temperatures, and one was frozen at -20°C to evaluate the effect of one freeze-thaw cycle. After 1, 3, 5 and 7 days, one aliquot of each experimental plasma was removed from 4°C, 30°C, and 37°C storage and processed as described for Day 0. The plasma aliquots frozen at -20°C on Day 0 were thawed and similarly processed on Day 8.

### HIV RNA extraction and reverse transcription

Each experimental plasma aliquot (140 μL) was manually extracted using the QIAamp Viral RNA Mini Kit (Qiagen, Valencia, CA) and eluted in 60 μL of elution buffer. Ten μL of each extracted RNA were reverse transcribed into cDNA using SuperScript IV (ThermoFisher Scientific, Waltham, MA) and random hexamers in a 20 μL reaction following the manufacturer’s protocol.

### HIV RNA quantification

The HIV cDNA from each experimental plasma aliquot was quantified in duplicate (10 μL/reaction) by a laboratory-developed qPCR targeting the HIV 5’ LTR [23]. Serial dilutions of 8E5 cell DNA [24] containing from 10^1^ to 10^4^ HIV copies/10uL were used as quantification standards.

### Data analysis

To define the intra-assay and inter-assay variation of the HIV qPCR assay, one standard deviation (**SD**) of the mean VL values measured for the C-h and the C-l were used, performed in a single assay and across three assays on separate dates, respectively. Consistent with published literature [25–28] a VL decline of >0.5 log_10_ c/mL was considered clinically relevant.

The effects of temperature on the decay of HIV RNA VL during the storage of experimental plasma were measured as mean log_10_ VL change of all 13 experimental plasmas from Day 0 to Day-1, D-3, Day-5 or Day-7, calculated separately and stratified by temperature. Lines of best fit for VL changes over time at each temperature were generated by linear regression, and the rates of change were compared to a slope of zero to assess statistical significance (GraphPad Prism, version 10). The effect of one freeze-thaw cycle on plasma HIV RNA VL was estimated by paired t-test. P-values of <0.05 were considered statistically significant.

## Results

### Variability of the quantitative HIV RNA qPCR

The intra-and inter-assay variability of our HIV RNA qPCR assay was <0.2 log_10_ c/mL, which is within the variability reported for most qPCR assays [29–31]. The SD for the controls quantified in a single assay were 0.06 log_10_ c/mL for C-H and 0.14 log_10_ c/mL for C-l, and the SD for controls quantified on three separate dates were 0.18 log_10_ c/mL for C-h and 0.11 log_10_ c/mL for C-l (**Table 1** in **S1 File**).

### Stability of HIV RNA in plasma held at various temperatures over time

The effect of one freeze-thaw was assessed by a comparison of the Day-0 and Day-8 VL of the 13 experimental plasmas held at -20°C which found a mean difference of -0.10 log_10_ c/mL (95% CI -0.26, 0.06; p=0.2) (**Tables 2** and **3** in **S1 File**).

Compared to Day-0, the mean VL for the experimental plasmas stored at 4°C and 30°C had minimal variation at Day-1, D-3, Day-5 and Day-7. At Day 7 the VL had a mean change of -0.01 log_10_ c/mL (95% CI -0.11, 0.09) and -0.08 log_10_ c/mL (95%CI -0.21, 0.05), respectively, both below the clinically relevant threshold of 0.5 log_10_ c/mL (**Fig 1** and **Table 3** in **S1 File**).

**Figure 1.**
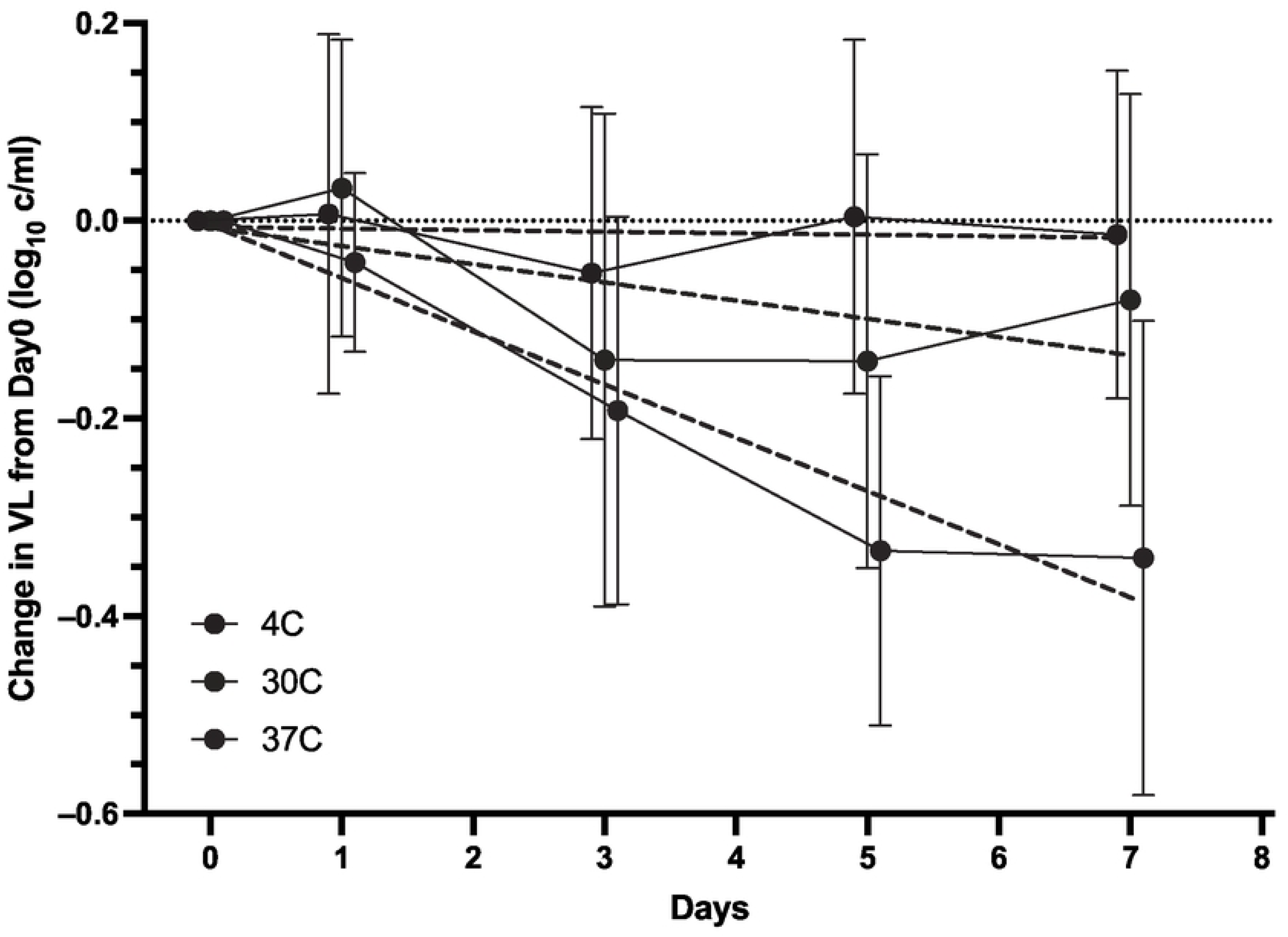
Effects of various temperatures above freezing for Day-1, Day-3, Day-5 or Day-7 on HIV RNA levels in plasma. Aliquots from 13 experimental plasma samples (HIV RNA VL range prior to storage (Day-0): 2.70-5.53 log_10_ c/mL) were stored at 4°C, 30°C and 37°C for 1, 3, 5 or 7 days prior to RNA extraction and quantification. The mean change in HIV RNA values from Day-0 (y-axis) among the 13 experimental plasmas stored at each temperature is plotted over time (x-axis). Each temperature is shown by a solid line and a unique color (4°C, 30°C or 37°C in blue, purple, and red, respectively). Standard deviations are indicated by whiskers. Linear regression (thicker dashed lines) was used to estimate plasma HIV RNA rates of decay over time at each temperature.

The mean change in the VL at Day-7 for the experimental plasmas stored at 37°C (-0.34 log_10_ c/mL, 95%CI -0.49, -0.20) was also <0.5 log_10_ c/mL; however, unlike the specimens stored at 4°C and 30°C, the specimens stored at 37°C demonstrated a progressive loss in VL: the mean changes observed from Day 0 to Day-1, D-3, Day-5 or Day-7 were -0.04, -0.19, -0.33. -0.34 log_10_ c/mL, respectively. Notably, three (23%) of the 13 specimens stored at 37°C had progressive VL losses over the week, so that by Day-7 the declines were greater than -0.5 log_10_ c/mL (S7= -0.67, S8= -0.78, S12= -0.50; **Table 3** in **S1 File**); these changes likely drove the observed progressive decay of the mean HIV RNA of plasmas stored at 37°C. These three experimental plasmas had Day-0 VL of 3.14, 3.59, and 4.19 log_10_ c/mL. In addition, we observed single plasma aliquots with sporadic HIV RNA losses greater than -0.5 log_10_ c/mL, but these specimens did not show progressive decay across time – these included three aliquots stored at 30° C (S1, S2, S6), and one stored at -20° C (S10). These were likely the result of technical artifacts and were not part of observed trends in RNA decay.

Linear regression across specimens stored at 4°C and 30°C found the average rates of VL decay over a period of 7 days were not clinically relevant nor statistically significant (slope -0.002, 95%CI -0.017, 0.014; p=0.77, and -0.018, 95%CI -0.057, 0.020; p=0.23, respectively). However, specimens stored at 37°C had a significant decline exceeding the clinically relevant threshold, with an average rate of decay of -0.054 (95% CI -0.079, -0.029; p=0.01) (**Fig 1** **and** **Table 4** in **S1 File**).

## Discussion

This study found that HIV RNA in plasma remained stable when stored at 4°C or 30°C for at least one week. Changes in HIV RNA levels were limited and similar to those observed after one freeze-thaw cycle on samples stored at -20°C.

However, when experimental plasma was stored at 37°C the HIV RNA progressively decayed compared to the values measured prior to storage. Importantly, among specimens stored at 37°C, significant declines in plasma HIV RNA, defined as a loss greater than 0.5 log_10_ c/mL, was most notable among specimens with VL of approximately 10^4^ log_10_ c/mL and lower.

Our finding of stable HIV RNA in samples stored at ≤30°C for one week compared to the HIV RNA values from these same plasmas prior to storage suggests that rural clinics with ambient temperatures ≤30°C could transport plasma without use of dry ice. Rather, transport of plasma to a centralized laboratory within a week while maintained at ambient temperatures ≤30°C would likely yield accurate plasma HIV RNA VL. The decay of HIV RNA in plasma stored for a week at 37°C indicates that RNA stability can be significantly compromised between 30°C and 37°C. This emphasizes that in warmer regions or in enclosed vehicles that capture heat, plasma specimens should be transported in insulated containers with cold packs that can maintain a temperature of ≤30°C. Maintaining plasma at ≤30°C is particularly relevant to the accurate diagnosis of low-level viremias given our findings that significant losses (more than -0.5 log_10_ c/mL) were observed in specimens stored at 37°C.

Our results are consistent with other studies that found HIV RNA was stable in plasma held at 4°C or 21°C for at least three days [18, 28, 32], and confirm two studies that found plasma HIV RNA stored at 4°C, 22°C and 30°C for one week did not significantly decay compared to fresh plasma using a significance threshold of a loss greater than -0.5 log_10_ c/mL [20, 33]. Of note, our study included four samples (31%) with VL near 1000 c/mL (three of these were below 1000 c/mL), thus confirming the stability of HIV RNA in plasma stored at ≤30°C in samples with low viremias. In contrast, another study observed that HIV RNA declined by 0.74 log_10_ c/mL after one week at 30°C [21]; however, in this study plasma was stored in lysis buffer, which disrupts the viral envelope and may have allowed RNA degradation.

Few prior studies have looked at stability of HIV RNA when plasma is held at temperatures above 30°C; yet many low-and middle-income countries commonly have temperatures above 30°C. In one study, the HIV RNA in samples stored at 37°C for one week had a median decrease of 0.92 log_10_ c/mL [20], while a second study found a median decrease of 0.42 log_10_ c/mL in plasma samples stored at 37°C for 6 days [34]. In these studies, only a subset of samples stored at 37°C, 3/10 and 2/5, respectively, had reductions in VL values greater than -0.5 log_10_ c/mL. In our study, we observed a progressive decay in VL that was statistically significant after one week at 37°C with a mean decrease in HIV RNA of -0.34 log_10_ c/mL (range = -0.78 - 0.13 log_10_ c/mL). Similar to observations in previous studies, only 3/13 specimens had declines in VL that exceeded 0.5 log_10_ c/mL (mean decay = -0.65 log_10_ c/mL), which as mentioned above, appeared to be greater in specimens with lower VL. The proportionately greater decay in samples with lower VL suggests that similar levels of RNases or other degradation pathways may degrade a relatively greater proportion of HIV RNA in plasma with lower VL when stored at elevated temperatures.

Changes in VL >0.5 log_10_ c/mL are generally considered clinically relevant [25–28] based on estimates of VL assay variability and biological variability within an individual [35]. In our study, as well as others, mean VL changes observed after one week of plasma storage at temperatures ≤ 30°C were below this threshold. Even when stored at 37°C, most plasma samples had declines of less than -0.5 log_10_ c/mL, except for a relatively small proportion of specimens that included some but not all plasma with the lowest VL. Whether the variability in VL observed after storage beyond current recommendations is clinically significant will depend on whether the original plasma viral load of a specimen falls close to the threshold used to define virologic failure. In one study, routine diagnostic plasma samples stored for up to a week at temperatures between 4°C and 30°C reliably differentiated between ART-suppressed and virologic failure (defined at ≥1000 c/mL) in 98.8% of 1194 cases [33].

Limitations of our study include the use of a laboratory developed quantitative HIV RNA qPCR instead of a commercial VL assay. This manual assay may have led to a small number of single plasma aliquots with sporadic HIV RNA losses >0.5 log_10_ c/mL, which were likely technical artifacts caused by performance of RNA extractions and cDNA reactions by different operators and on different dates when environmental conditions may vary. Use of automation for extractions and assay set-up would have likely mitigated these occasional variable results. Nonetheless, using laboratory controls we showed that the intra- and inter-assay variability of our HIV LTR qPCR assay was within the variability reported for most qPCR assays [29, 30][31]. Another study limitation is the storage of experimental plasma samples in a temperature-controlled laboratory setting that may not reflect the effects of temperature fluctuations that can occur in real world settings. Additionally, while the experimental plasmas were studied for a maximum of 7 days, a longer time may be required for specimen transport from some rural low-resource settings. However, our goal was to look at decay within the optimal timeframes for returning VL data to clinicians for patient management.

In conclusion, our study adds precision around the stability and decay of HIV RNA in plasma held at the ambient temperature of most laboratories. We provide temperature and time parameters for storage and transport of plasma to a centralized laboratory without compromising the VL results. Short-term storage and transport of plasma at ambient temperatures would greatly simplify logistics, reduce shipping costs and facilitate frequent shipping from rural clinics that are required to use centralized laboratories to assess VL. More rapid turn-around- times for VL measurements would allow timely management of low level viremias and virologic failure, which should improve treatment outcomes of HIV infected individuals and reduce the spread of HIV infection in resource-limited settings [17].

## Supporting information

**S1 File**. **The S1 File includes Tables 1, 2, 3 and 4.**

## Data Availability

All data associated with this study are available in the main text and Supporting Information.

